# Diffusion model enables quantitative CBF analysis of Alzheimer’s Disease

**DOI:** 10.1101/2024.07.01.24309791

**Authors:** Qinyang Shou, Steven Cen, Nan-kuei Chen, John M Ringman, Junhao Wen, Hosung Kim, Danny JJ Wang, Alzheimer’s Disease Neuroimaging Initiative

**Affiliations:** Laboratory of Functional MRI Technology (LOFT), Stevens Neuroimaging and Informatics Institute, University of Southern California, Los Angeles, CA, United States; Department of Biomedical Engineering, University of Arizona; Department of Neurology, Keck School of Medicine, University of Southern California, Los Angeles, CA, United States; Laboratory of AI & Biomedical Science (LABS), Stevens Neuroimaging and Informatics Institute, University of Southern California, Los Angeles, CA, United States; Laboratory of Neuro Imaging (LONI), Stevens Neuroimaging and Informatics Institute, University of Southern California, Los Angeles, CA, United States

## Abstract

**Objectives:** Cerebral blood flow (CBF) measured by arterial spin labeling (ASL) is a promising biomarker for Alzheimer’s Disease (AD). ASL data from multiple vendors were included in the Alzheimer’s Disease Neuroimaging Initiative (ADNI) dataset. However, the M0 images were missing in Siemens ASL data, prohibiting CBF quantification. Here, we utilized a generative diffusion model to impute the missing M0 and validated generated CBF data with acquired data from GE.

**Methods:** A conditional latent diffusion model was trained to generate the M0 image and validate it on an in-house dataset (*N*=55) based on image similarity metrics, accuracy of CBF quantification, and consistency with the physical model. This model was then applied to the ADNI dataset (Siemens: *N*=211) to impute the missing M0 for CBF calculation. We further compared the imputed data (Siemens) and acquired data (GE) regarding regional CBF differences by AD stages, their classification accuracy for AD prediction, and CBF trajectory slopes estimated by a mixed effect model.

**Results:** The trained diffusion model generated the M0 image with high fidelity (Structural similarity index, SSIM=0.924±0.019; peak signal-to-noise ratio, PSNR=33.348±1.831) and caused minimal bias in CBF values (mean difference in whole brain is 1.07±2.12ml/100g/min). Both generated and acquired CBF data showed similar differentiation patterns by AD stages, similar classification performance, and decreasing slopes with AD progression in specific AD-related regions. Generated CBF data also improved accuracy in classifying AD stages compared to qualitative perfusion data.

**Interpretation/Conclusion:** This study shows the potential of diffusion models for imputing missing modalities for large-scale studies of CBF variation with AD.

## INTRODUCTION

Alzheimer’s Disease (AD) is a progressive and devastating neurological condition that affects millions of individuals. Neuroimage biomarkers for the early detection of AD and/or mild cognitive impairment (MCI) play an important role in guiding preventative intervention and/or anti-amyloid treatment for potential progression of AD[1], [2], [3], [4]. Arterial Spin Labeling (ASL) is a non-invasive method to quantitatively measure cerebral blood flow (CBF)[5]. It has been shown that ASL CBF is highly consistent with fluorodeoxyglucose positron emission tomography (FDG-PET)[6], which is one of the current clinical standards for AD diagnosis. A recent systematic review[7] summarizing 81 original studies reported widespread CBF reductions, particularly in temporoparietal and posterior cingulate regions in AD individuals; MCI was also associated with decreased CBF in the posterior cingulum. Further, the patterns of ASL CBF reductions are correlated with disease severity and progression of AD[8]. This demonstrates the potential use of ASL CBF as an imaging biomarker for the early diagnosis and prognosis of AD[9].

Alzheimer’s Disease Neuroimaging Initiative (ADNI) (https://adni.loni.usc.edu/) is one of the pioneering open-access datasets for studying AD that includes ASL in the research protocol. In particular, the ADNI-3 study includes 3D ASL data from three major MR vendors, offering the opportunity to illustrate quantitative CBF evolutions with AD progression. However, calculating CBF requires both perfusion-weighted images and a proton density or M0 image. Currently, the implementation of ASL techniques is different across vendors. For example, General Electric (GE) uses pseudo-continuous ASL (pCASL) with M0 for CBF quantification. In contrast, 3D ASL data acquired on Siemens scanners, which consist of >50% of the ADNI-3 dataset, used pulsed ASL (PASL) without the M0 image, prohibiting quantitative CBF calculation. This absence of M0 images in ADNI-3 dataset greatly impairs the potential of using CBF as a biomarker for AD.

Deep-learning (DL) based generative models such as generative adversarial networks (GANs) and variational auto-encoders (VAEs) have made remarkable progress in the past few years, including synthesizing realistic natural images[10], [11], [12], [13], and biomedicine[14], [15]. Recently, generative diffusion models[16] have been developed and shown the ability to generate high-quality images[17], [18]. In principle, a diffusion model generates an image from random noise through a series of iterative, denoising steps guided by the learned probability distribution. It has also been used to perform cross-modality image-to-image translation [19]. For example, Lyu et al. developed a diffusion model for conversion between CT and MRI images[20]. In another study, Meng et al. used a score-based model to learn from multimodal image data and generate missing imaging modalities for dataset completion on the BraTS19 dataset[21]. These works have shown the feasibility of using diffusion models for generating high-fidelity medical images. The latent diffusion model (LDM)[22] is a recently introduced technique that performs a diffusion process in the latent space to improve computational efficiency while providing a flexible conditioning mechanism. So far, there is still limited evidence to support using these DL-generated images in neuroimaging studies, such as the ADNI.

In this work, we developed a conditional LDM to generate the M0 images for Siemens ASL data in ADNI-3 to quantify CBF and hypothesized that the pattern of decreasing CBF with AD progression could be observed in the generated data. Figure 1a illustrates the architecture of our proposed LDM model, and Figure 1b shows the workflow of the study evaluation. We first demonstrated the feasibility of using conditional LDM to generate M0 images with high fidelity and accuracy for CBF quantification on our in-house dataset (see Supplementary Table 1 and Method section for details). Then, we showed the feasibility of applying the trained model to the Siemens ASL dataset in ADNI-3 to impute the missing M0 images for CBF quantification. Further, we compared the patterns of CBF variation with AD progression from generated (Siemens) and acquired (GE) CBF data with regional analyses. Three machine learning classifiers were applied to evaluate the capability of CBF and perfusion-weighted images to classify different AD stages. Our work demonstrates that the proposed diffusion model can impute missing data modality and provide sufficient power and precision for studying ASL CBF as a biomarker in AD.

**Figure 1.**
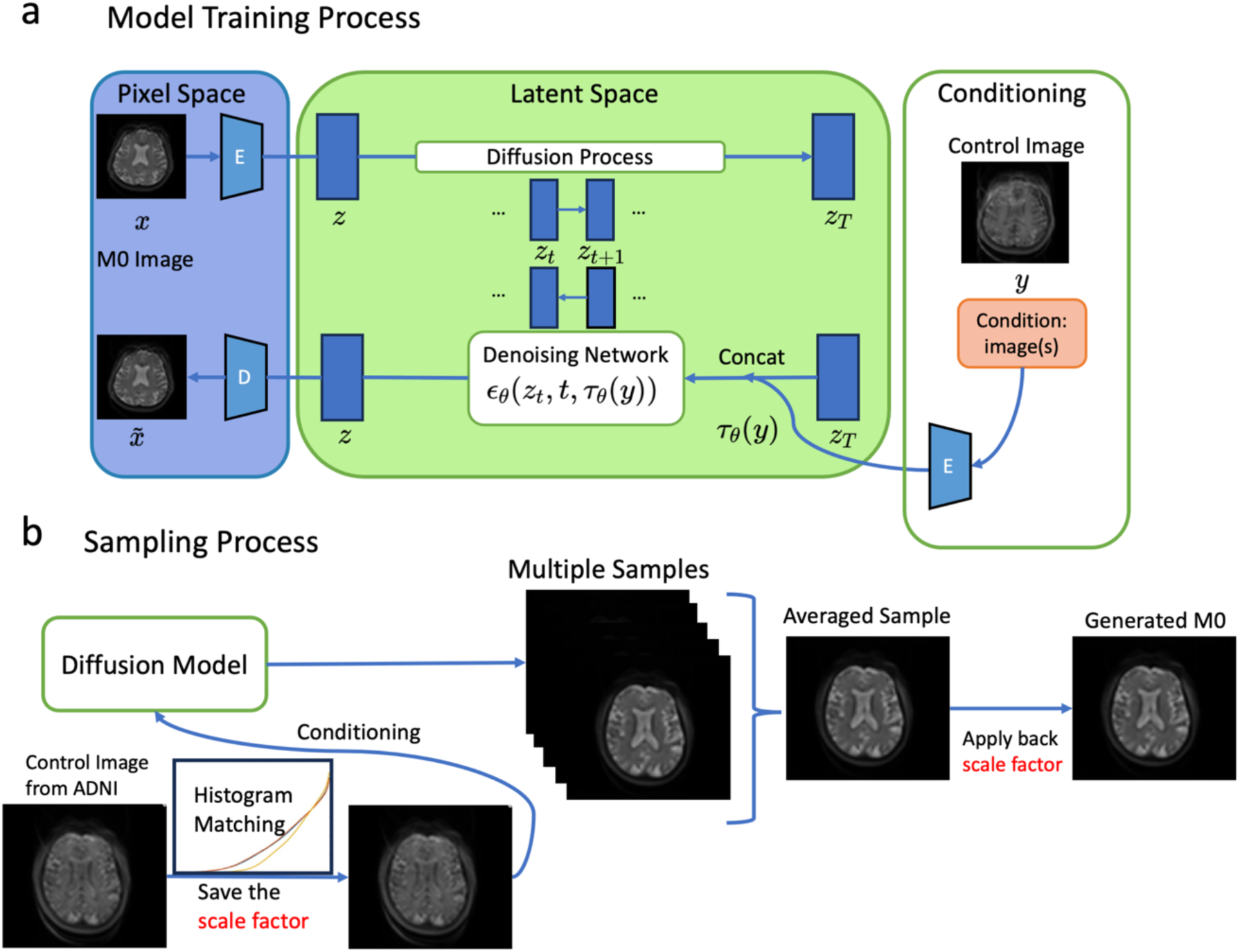
Illustration of the conditional latent diffusion model(LDM) and the inference process. a. Model structure of the latent diffusion model. The image (M0) in the original pixel space is first encoded into the latent space with the encoder(E). Diffusion process is conducted in the latent space to improve the computational efficiency. The condition (control image) is also encoded into the latent space with the same encoder and concatenate with the latent space feature of M0. The reversed denoising network *∈_θ_* takes the time step *t*, latent space image *z_t_* and the latent-space image of the condition *τ_θ_*(*y*) as input and produces the previous latent space image in the previous step. The final M0 image is produced with a decoder to recover the image from the latent space. b. The sampling process (generation). The input images are first scaled with the histogram matching to match the histogram of the training data. The trained LDM generate 20 samples for each scan and averaged to get a robust sample. The scale factor saved in the previous histogram matching is used to scale the image back to the original scale.

## RESULTS

### Testing conditional latent diffusion model on an in-house dataset

We first trained and validated the conditional LDM on our in-house dataset (N=55, age=73± 7, 40 females) with similar age as the ADNI cohort (see Supplementary Table S1 and S2). Ground truth M0 images were acquired by manually turning off the background suppression pulses and applying a long inversion time (TI=5 sec) in PASL acquisitions with a Siemens scanner. Since the LDM employs probabilistic models, there are variations among the generated M0 images. Therefore, we generated multiple M0 images followed by averaging and plotted the curves of performance metrics as a function of the number of averaged images, as shown in Figure 2a, b, and c for normalized mean squared error (NMSE), structural similarity index (SSIM) and peak signal-to-noise ratio (PSNR) between the generated M0 and the ground truth (acquired M0 image), respectively. The quality of the averaged M0 image improved with an increasing number of averages and got stabilized after averaging 20 samples. The variation of performance metrics across all test subjects (indicated by the shaded area) illustrates the stability of the conditional LDM across different subjects. Figure 2d, e, and f show one representative case in the test dataset for an averaged M0 image, the ground truth, and the difference map between the two images, respectively. Figure 2g shows the standard deviation map across 100 generated samples. Most brain regions have low standard deviation, supporting the high certainty of the trained conditional LDM. The regions with relatively higher standard deviations correspond to cerebrospinal fluid (CSF) or vasculature, as red arrows indicate, which tend to have high fluctuations in the MR images.

**Figure 2.**
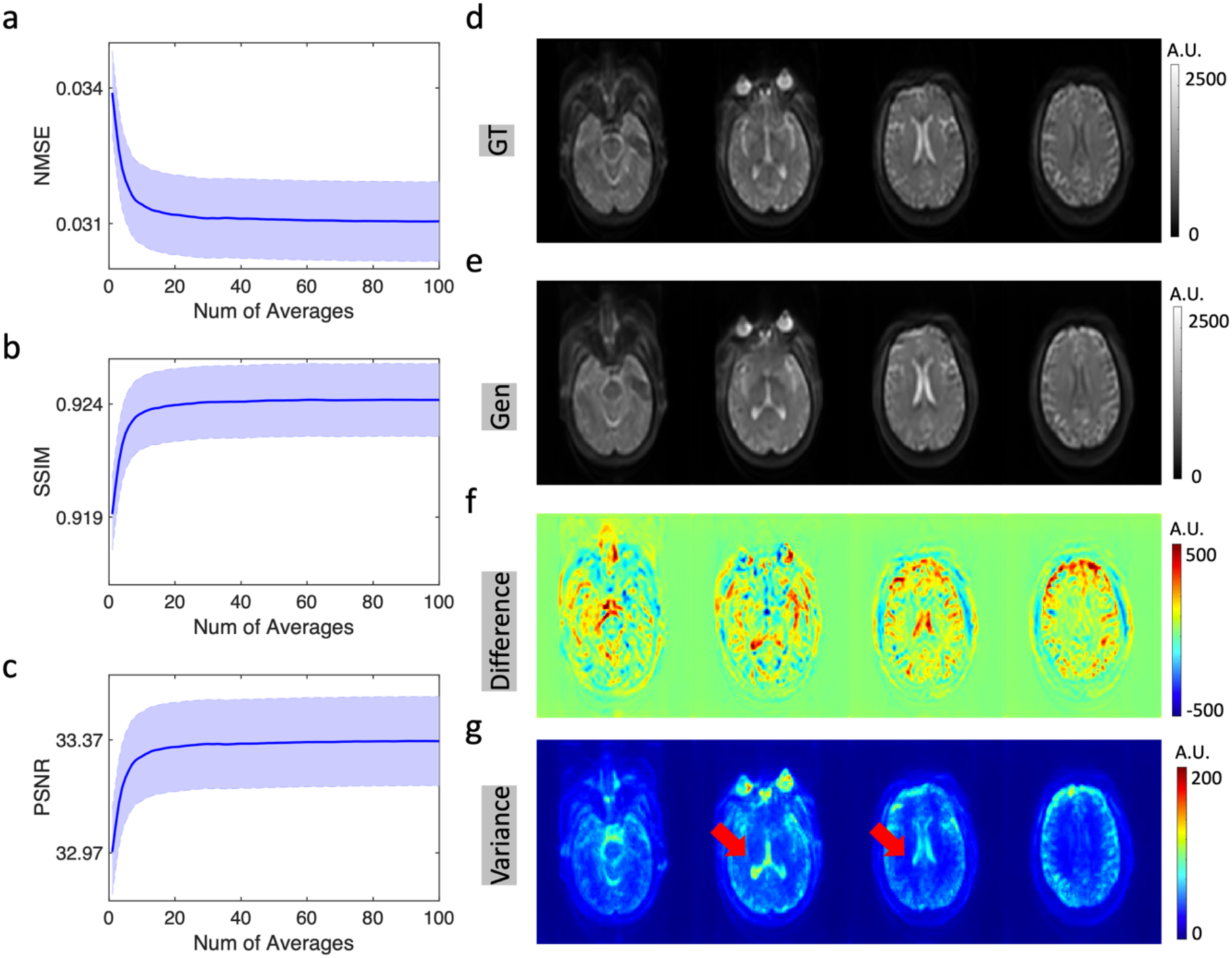
Uncertainty evaluation of the diffusion model. a, b and c. Normalized mean square error (NMSE), structural similarity index (SSIM) and peak signal-to-noise ratio (PSNR) between generated and ground truth M0 image with different number of averaged samples. The shaded area shows standard deviation across test images. d and e. A representative case of the ground truth (GT) and the generated image. f. The difference map between the ground truth and the generated M0 image. g. Standard deviation map across 100 generated samples. Red arrows show CSF areas where the standard deviation is higher than other brain tissues.

Based on the results of our uncertainty evaluation, we chose to average 20 generated images for each case in our following analysis to achieve the optimal balance between generation time and image quality. We systematically compared the generated M0 and CBF maps to the ground truth images in the following aspects. First, a representative case of generated M0 and CBF images are shown in Figure 3a, both of which have high similarities to the ground truth. Second, the quantitative similarity metrics of the generated images are summarized in Figure 3b, demonstrating the excellent performance of the conditional LDM. The mean gray matter (GM) and white matter (WM) CBF values are further plotted between generated CBF maps and the ground truths, showing a high consistency (r=0.97) between the generated and true CBF values. Finally, Figure 3d shows the mean difference between the generated CBF values and the ground truths in the whole brain, GM, and WM across all test cases. The mean difference is 1.07±2.12ml/100g/min for the whole brain, 1.29±2.51ml/100g/min for GM, and −0.04±1.44ml/100g/min for WM, which is less than 5% of the corresponding CBF values. This shows our model’s feasibility and fidelity in preserving the qualitative and quantitative features of the M0 image.

**Figure 3.**
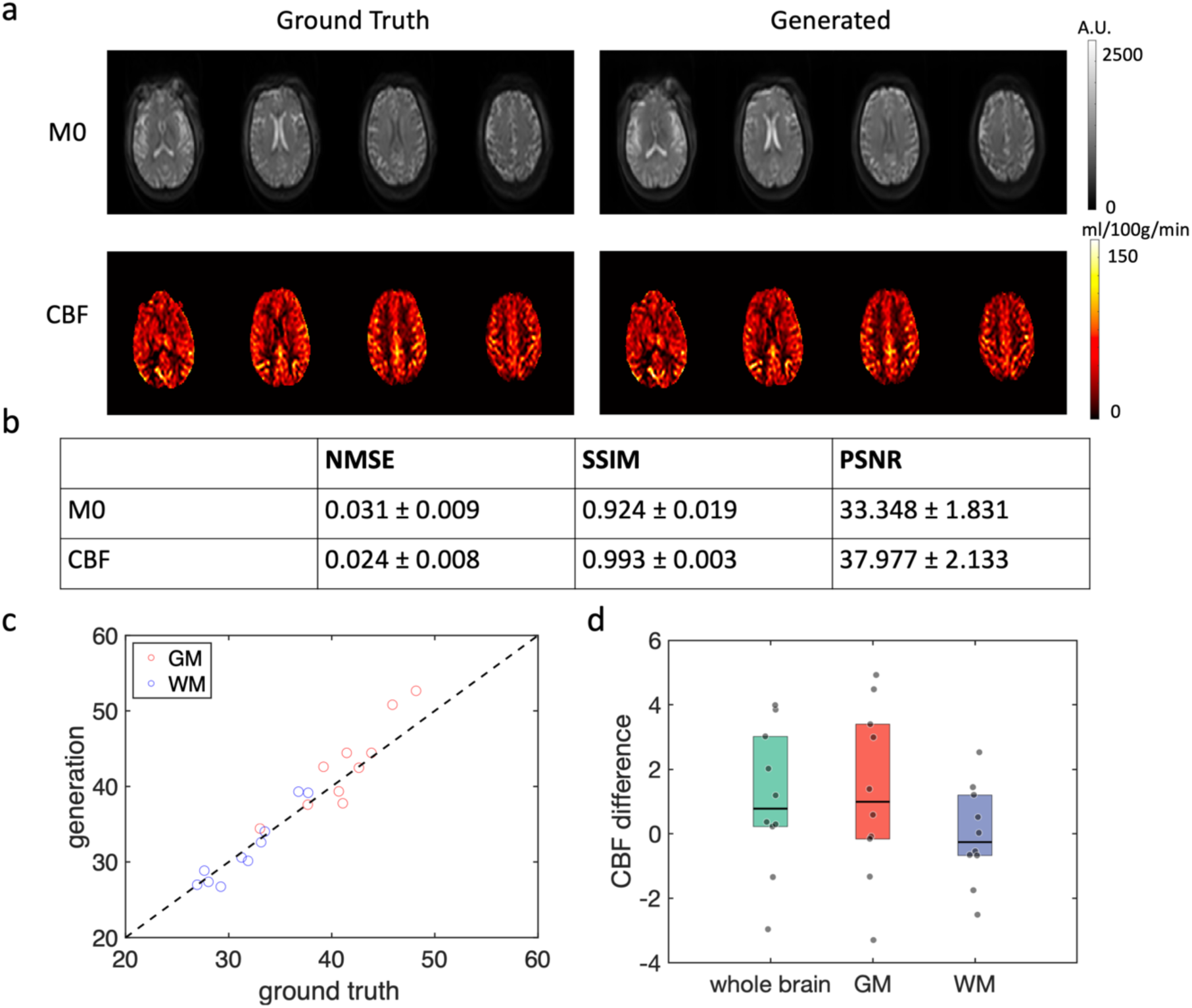
Qualitative and quantitative model performance evaluation. a. A representative case of the ground truth and generated M0 and CBF maps. Both M0 and CBF map produced by the diffusion model show high fidelity and similarity to the ground truth image. b. Quantitative similarity metrics of the generated image and the ground truth including NMSE, SSIM and PSNR. c and d. Bias in CBF quantification measurements. c. Scatter plot of the averaged CBF values of the ground truth and generated CBF images in both gray matter (GM) and white matter (WM). d. Averaged CBF difference in the whole brain, GM and WM. Mean difference is 1.07±2.12ml/100g/min for whole brain, 1.29±2.51ml/100g/min for GM and −0.04±1.44ml/100g/min for WM.

We further evaluated the fidelity of our generated M0 images based on their consistency with the MR physics model. The relationship between M0 and the control image is dependent on the timing of background suppression (BS) pulses, as shown in Supplementary Figures S1 and S2, as well as expressed by Equation 3 in the Method. We calculated the T1 map from the generated M0 image and the acquired control image and compared that with the ground truth and the reported T1 values of normal brain tissue at 3T in the literature. Supplementary Figure S2c and d show a high similarity between the T1 map calculated from the generated M0 and that calculated from the acquired M0. The largest variation between the generated T1 map and the ground truth T1 map occurs in the area containing blood vessels and CSF, where the signal fluctuation is relatively large. Supplementary Figure S2e and f show the averaged T1 values of the ground truth and fitted from the generated M0. T1 values fitted from the generated M0 are close to the ground truth, and T1 values of both GM and WM are consistent with that reported in literature[23], confirming that our generative model preserves the MR physics properties.

### Evaluation of the diffusion model on the ADNI dataset

Since the diffusion model was trained on our in-house dataset acquired with the same vendor, and the imaging parameters are identical to those of the ADNI dataset, the acquired images were expected to have similar image contrast. Therefore, we directly applied the trained model to generate the missing M0 images for the Siemens ASL dataset in ADNI-3. ADNI-3 acquired ASL data with 10 pairs of control/label images for each scan, the average control image was used as the condition to generate the M0 image. Intensity normalization was performed to make the intensity of the control images from the ADNI dataset match with that of the in-house dataset so that the model could achieve optimal performance. Like the previous steps, 20 samples were generated for each scan and averaged to produce the final M0 image for CBF calculation. CBF maps were calculated with the acquired perfusion images and the generated M0 images according to Equation (1) (in Materials and Methods section). The demographic information of the ADNI dataset is summarized in Supplementary Table S2. All subjects were characterized into four diagnostic groups, including cognitive normal (CN), subjective memory concerns (SMC), mild cognitive impairment (MCI), and AD at each visit. To compare the spatial patterns among the four groups, averaged CBF maps were calculated for each group. Both averaged perfusion maps (i.e., a direct subtraction between control and label images) and CBF maps were calculated and compared to justify the need for quantitative analysis. Figure 4a-d show the averaged perfusion and CBF maps of each group for both Siemens and GE data, respectively. The perfusion maps are in arbitrary units, while the CBF maps are standardized to the unit of ml/100g/min. A greater degree of spatial inhomogeneity can be observed in the perfusion maps compared to CBF maps, especially in top and bottom slices, which is likely due to spatial variations of coil sensitivities and different head positions. The CBF maps show improved homogeneity by normalizing the perfusion signal with a calibration scan (M0).

**Figure 4.**
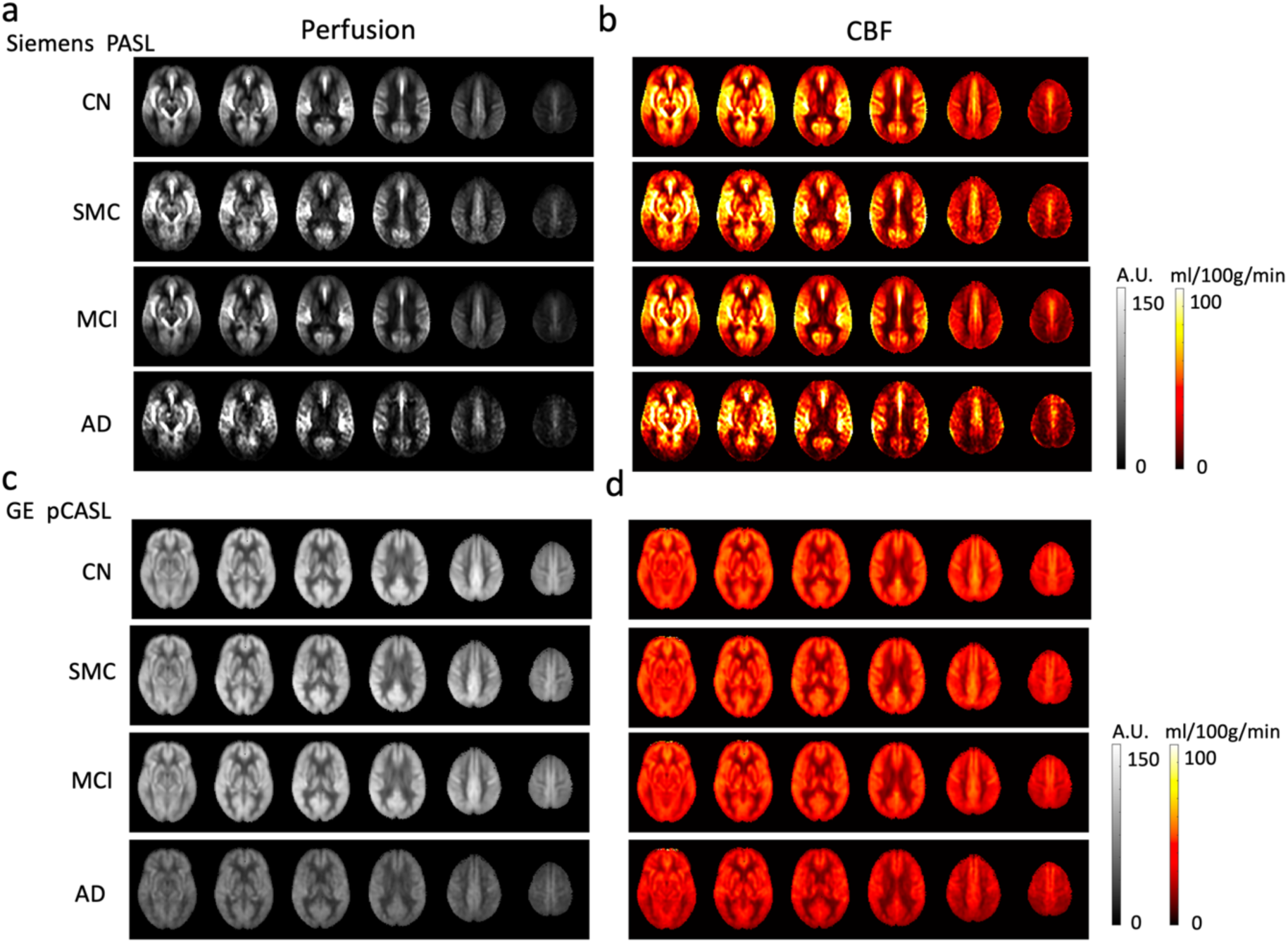
Averaged perfusion and CBF maps for different groups in the ADNI dataset including CN, SMC, MCI and AD. The perfusion maps show larger inhomogeneity across different brain regions, while CBF maps are more homogeneous. Both Siemens and GE data show a decreasing trend from CN/SMC to MCI to AD, especially in the CBF maps. Siemens pulsed ASL data show more vascular signals while GE pCASL data is smoother across the whole brain.

### Results of Regional Analysis

The perfusion and CBF maps were normalized to the canonical space of the MNI brain template according to their corresponding T1 images. Mean CBF and perfusion values in the 90 regions of interest (ROIs) defined by the Automated anatomical labeling (AAL) template[24] were extracted for further analyses.

We first compared the regional CBF values of baseline visits in ADNI-3 between generated data (Siemens) and acquired data (GE) across the four diagnostic groups using a generalized linear model with interaction between the diagnosis group and scanner types. The interaction test showed in 84 out of 90 ROIs, there are no statistically significant differences in CBF variation across NC, SMC, MCI and AD groups between generated (Siemens) and acquired (GE) data. We did not adjust for multiple comparisons to prevent false claims of no difference by penalizing α level. Details of the ROI results can be found in Supplementary Table S3. Based on the latest systematic review[7], AD is most affected by temporoparietal and posterior cingulate cortex (PCC) regions. Figure 5 shows boxplots of CBF values among the four groups in 4 representative ROIs, including PCC, precuneus, angular gyrus, and inferior temporal gyrus for both left and right hemispheres. Boxplots of CBF values in 4 other AD-affected regions (cuneus, inferior parietal gyrus, supramarginal gyrus, and hippocampus on both hemispheres) are shown in Supplementary Figure S3. GE and Siemens data are shown in red and blue, respectively. Visual inspection concurred with the interaction test and showed a high level of similarity between generated and acquired CBF data, with a trend of decreasing CBF from CN to MCI and AD, while there is little CBF difference between CN and SMC. Siemens data had greater variances than GE data, as shown by larger interquartile ranges in the box plot.

**Figure 5.**
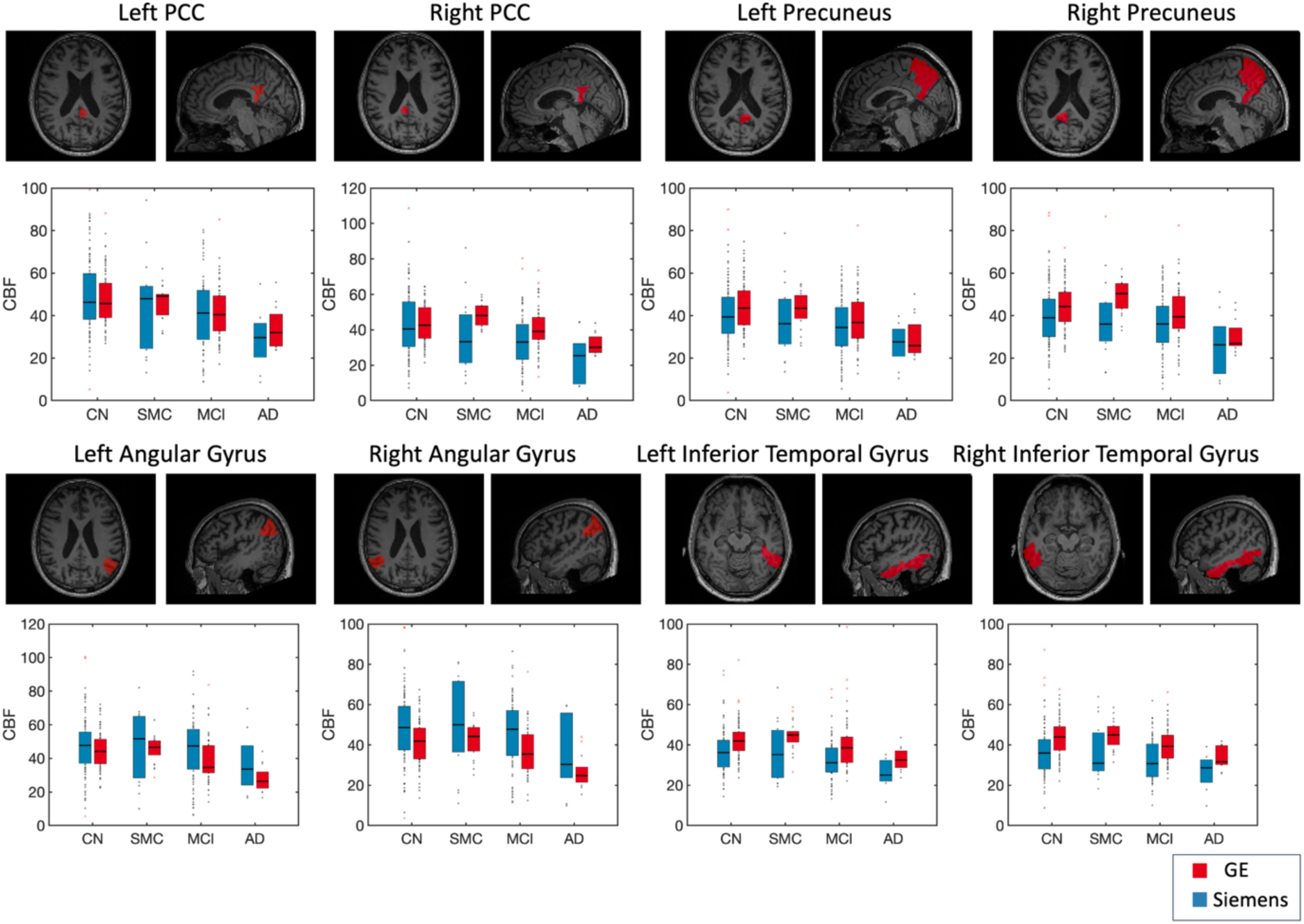
Regional analysis of the trend in different groups of the subjects. Four AD-related ROIs are shown including posterior cingulate cortex (PCC), precuneus, inferior parietal gyrus and angular gyrus. Data from Siemens and GE are shown in blue and red respectively. Both data show similar trend in these ROIs, with a decreasing from CN/SMC to MCI to AD.

### Results of Machine Learning Classification

To study whether using quantitative CBF data can improve the performance of binary classification between each pair of the 4 diagnostic groups than using qualitative perfusion data, three machine learning (ML) methods, including Ada Boost[25], Elastic Net and Random Forest, were trained with regional CBF and perfusion values as features respectively. The performances of ML classifiers are shown in Figure 6 and Supplementary Table S4. The receiver operating characteristic (ROC) curve of three ML methods for 4 pairwise group classifications, including AD vs CN, AD vs MCI, MCI vs CN, and SMC vs CN are displayed. For both Siemens and GE data, the performance is above acceptable for clinical use[26] to separate AD from CN, with the best area-under-the-curve (AUC) of 0.75 95% CI: (0.62, 0.89) for Siemens data and 0.9 95% CI: (0.83, 0.98) for GE data, while the performance for classifying MCI and CN is below the clinically acceptable level of 0.7 for both Siemens and GE. The performance using perfusion features is lower than that using quantitative CBF features in most cases except for MCI vs CN, supporting the importance of using quantitative CBF rather than qualitative perfusion values for classification.

**Figure 6.**
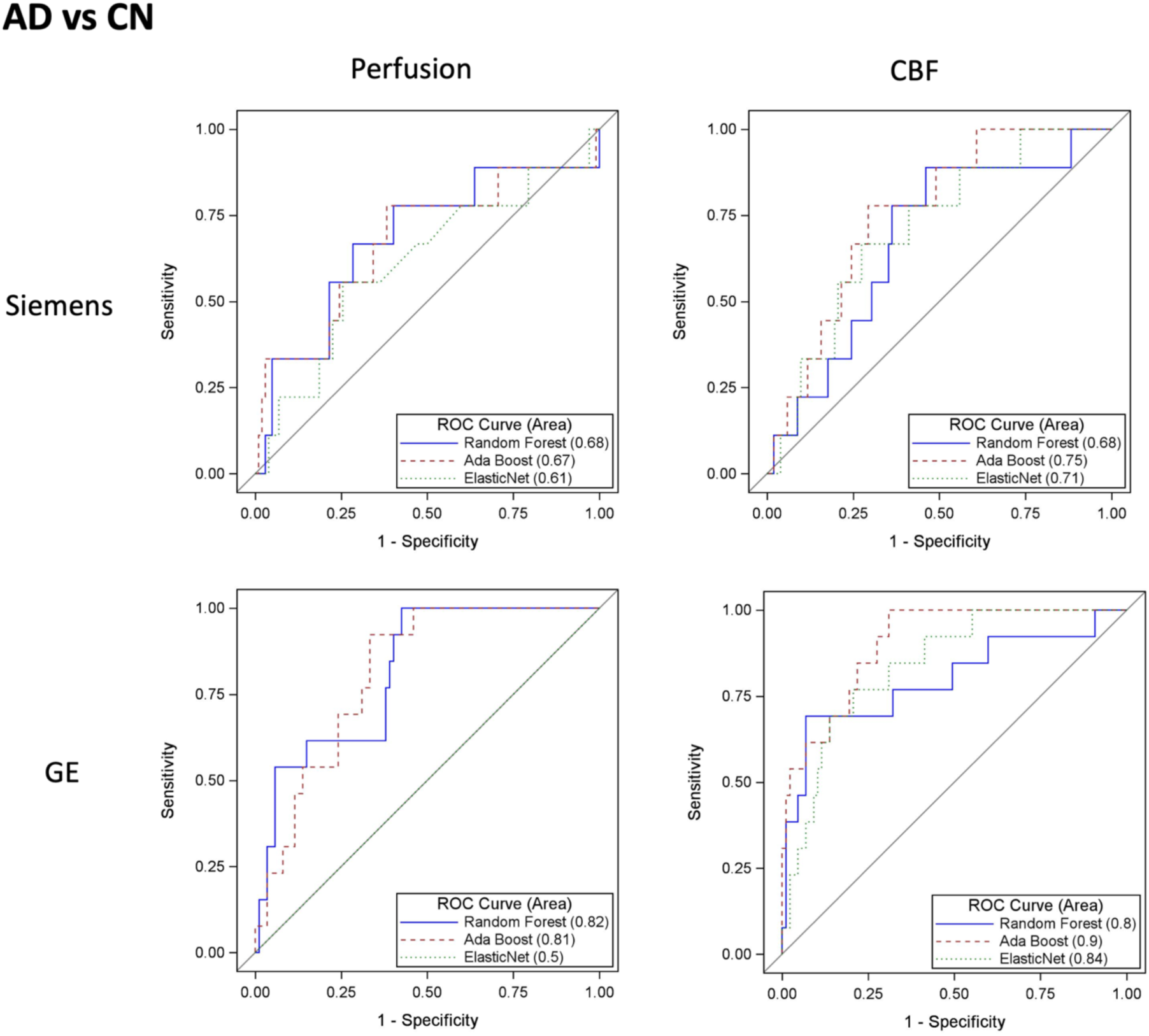
Performance of machine learning classification between AD and CN. ROC curve of the classification from three machine learning methods including Ada Boost, Elastic Net and Random Forest are plotted. For both data from GE and Siemens, the performance of classification is better when using CBF used as features compared to using perfusion as features. The results from Siemens data are slightly lower than GE data.

### Longitudinal Analysis of CBF Trajectories

We further explored longitudinal analyses on participants with multiple visits in ADNI-3 using a mixed effect model using an autoregressive covariance structure. The details of the sample size with different numbers of follow-ups are listed in Supplementary Table S5. The trends of CBF progression from AD related regions by diagnostic groups and scanner types are displayed in Supplemental Figure S5. The slope difference with a 95% confidence interval was estimated by a mixed model with a post hoc contrast test (results shown in Supplementary Table S6). In 39 out of 90 ROIs, there is no significant difference in slopes in any of the 4 groups between Siemens and GE. In 35 of 90 ROIs, there is significant difference in slopes in only one of the 4 groups between Siemens and GE.

## DISCUSSION

In this work, we demonstrated that the generative diffusion model trained on our in-house dataset can be successfully applied to an existing public dataset (ADNI-3) to generate M0 images conditioned on the control image for PASL acquisition. With the conditional LDM, we can achieve high-quality and high-fidelity M0 images without systematic bias in CBF quantification while preserving the consistency with the MR physics model. We also showed that the generated M0 image could be used for quantification of CBF and prediction of CBF variations in relation to AD progression, which has a better performance compared to non-quantitative perfusion analyses. The patterns extracted from the generated CBF maps were validated by comparing with ASL dataset acquired from another vendor, which were consistent with the literature. It also provides insights for generating other missing modalities for existing public datasets for multi-model studies.

ASL has been incorporated in many studies of neurological diseases[27], [28]. The two main advantages of using ASL to measure brain perfusion are its non-invasive nature and its quantitative capabilities. These characteristics make ASL potentially generalizable across different scan protocols, vendors, and sites. However, M0 is necessary to achieve quantitative CBF measurements. The original perfusion images can vary from site to site, even with identical imaging protocols, due to different settings of the scanner hardware and/or software. Such variations may affect the results of group analyses with multi-site data because the variance might arise from different sites rather than different diagnostic groups. Our study shows that the conditional LDM can generate the missing M0 image from the acquired control image without causing bias in quantification, and these generated images are particularly valuable for further quantitative analysis together with the existing data. This method can provide a robust method to recover those missing modalities in existing datasets, facilitating the analysis of multi-site ASL and other MRI data.

Recent developments in DL and generative models make it possible to perform image-to-image translation in many different ways. For example, this can be achieved by training a neural network to directly map the input image to the desired output or using conditional GANs to generate a new modality from the input[29]. Compared to these methods, using the diffusion model offers advantages such as better interpretability, shown by the uncertainty maps, and better flexibility to add conditions given its mechanism of using conditional probability propagation. In this work, we chose to generate the M0 image first and calculate the CBF map with the known kinetic model instead of directly generating the CBF image by using the perfusion and control image as the condition. This is because the SNR of the perfusion image is generally much lower than the control and M0 images, which indicates there are larger variations in the perfusion images, making the model less stable. This is confirmed with our preliminary study[30], where the model that generated the M0 image followed by CBF calculation yielded better performance than the model that directly generated the CBF map. Furthermore, the step-by-step nature of this workflow enhanced interpretability for generating CBF maps.

In our study, we observed similar patterns of CBF reduction associated with AD progression from generated CBF data, which was consistent with previous studies[8], [9], and acquired ASL data from another vendor. However, there were still differences in regional CBF values between acquired GE and generated Siemens data, with the latter demonstrating larger variations across subjects. This is likely due to the difference between pCASL (GE) and PASL (Siemens) data. In general, with the identical inversion time (TI) and post-labeling delay (PLD), pCASL yields higher SNR compared to PASL as more blood has reached the imaging slice at the time of acquisition[31], which explains interindividual variations in PASL data. In addition, a greater amount of arterial transit effects indicated by intravascular signals can be observed in PASL images around the Circle of Willis and its major branches. The difference between pCASL and PASL protocols may also lead to different quantitative CBF values, although the overall CBF trends with AD progression are comparable between Siemens and GE data.

Traditional machine learning approaches are shown to have comparable performance with more complex DL methods for classification [32]. In our studies, ML achieved overall good performance to classify AD from CN. The ML results showed reduced but acceptable accuracy in classifying AD from NC using generated data compared to the acquired data. This could be due to the small sample size (N=9) of AD patients. For the performance from larger sample categories, e.g., MCI vs. NC, the performance gap is closer. Since the comparison between generated and acquired images was not based on the same cohorts, the comparison could be confounded by demographic factors and comorbidities as well as different ASL techniques used.

There are some limitations of this study. First, there is no ground truth to be used as the reference for the generated M0 images in the ADNI-3 Siemens dataset. In this study, we evaluated the fidelity of the proposed method by several validation methods, such as validating the T1 maps with the physics model and literature, as well as by comparing the detected patterns of CBF reductions across different groups between generated and acquired CBF data. Although this cannot completely replace the necessity of the ground truth image, this approach is suitable for real-world scenarios where the dataset has already been acquired. Future studies could utilize datasets with existing references to validate further the use of generative models in large multi-site datasets. Second, pCASL data has higher SNR than PASL, which is also indicated in our results, where GE pCASL data showed better performance in classification than Siemens PASL data. Future studies should consider this difference and further standardize the imaging protocol of ASL. Third, the ADNI dataset is imbalanced, with more CN and MCI individuals than AD, which may affect the statistical significance of group comparisons and the performance of ML classifications. Fourth, the comparison between Siemens and GE was based on different patient cohorts, which could be confounded by demographic factors and comorbidities.

## MATERIALS AND METHODS

### Components of CBF quantification

ASL is an MRI technique that uses inversion pulses to label the water in the blood so that the perfusion weighted signal can be measured by subtracting the labeled image from the unlabeled, or the control image. The quantification of CBF with two different type of ASL, namely PASL and pCASL, are described with the following equations:

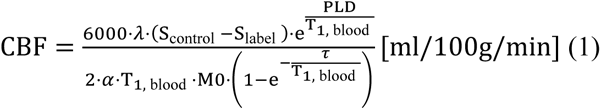

for pCASL and

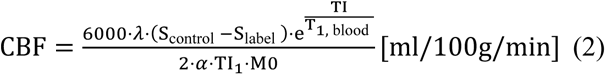

for PASL. Both equations require two key components, the perfusion signal, denoted by S_control_ − S_label_ and the proton density image, M0. M0 can be considered as a calibration image, which makes the CBF quantification robust against variations of imaging protocols, coil sensitivities, etc.

### Relationship between M0 and PASL

The sequence diagrams for PASL control/label and M0 are shown in Supplement Figure S1(a) and (b). Control and label images are acquired with two background suppression (BS) pulses in order to improve the SNR and motion robustness of the perfusion signal[29]. M0 is acquired without BS pulses and with a prolonged TI of 5 sec to enable full recovery of longitudinal magnetization. For a fixed timing of the BS pulses, the ratio between the intensities of the control and M0 image can be described as a function of tissue T1 and the inversion efficiency of the BS pulses.

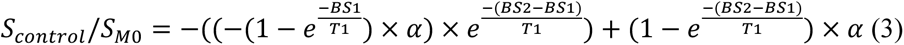

where *α* denotes the inversion efficiency of the inversion pulse for BS. However, without an explicit measurement of the values of tissue T1, such as T1 mapping, it cannot be directly derived from the control image. In this sense, there is no analytical solution to derive the M0 from the control image. The simulation of the control signal with different T1 values is shown in Supplement Figure S2(a) and (b). It can be seen that the control/M0 ratio is stable across different inversion efficiencies for most of the range of tissue T1s.

### Latent diffusion model

The conditional latent diffusion model in this study is illustrated in Figure 1a. It is consisted of an encoder, a diffusion module in the latent space, and a decoder. Prior to the forward diffusion process, the input image (M0) is first encoded into the latent space to reduce image dimension while not losing the perceptual content of the original image. Random Gaussian noise is added to the latent-space image for T steps (T=1000 in this study) to generate the intermediate representations until the data is transformed to a sample that follows Gaussian distribution. The reverse denoising process is modelled using a trainable network to iteratively recover the latent-space image in the previous step from these intermediate representations. The final output of the reverse diffusion process is fed to the decoder to convert the latent-space image to the image in the original space. This encoder-decoder pair was pretrained as in[22] and the model weights were directly adopted in this study. The conditioning mechanism is shown on the right side of Figure 1(a). The condition (control image) is first encoded with the same encoder and its representation in the latent space is concatenated with the latent-space image in the intermediate representations of M0 in each step of the reversed diffusion process to perform the conditional probability propagation.

### In-house dataset for model training and testing

The details of the imaging parameters of ADNI-3 Siemens PASL include TR=4000ms, TE=20.26ms, segmented 3D GRASE readout with a matrix size of 128×128×32 and a resolution of 1.9×1.9×4.5mm^3^, TI=2000ms and TI_1_=800ms. Two BS pulses were applied after the PASL inversion pulse to suppress the control/label images. In order to build an M0 generation model from the control images, we collected new dataset with paired data of 1)PASL with the same protocol as the ADNI-3 Siemens protocol, and 2) manually disabled BS pulses and extended inversion time (TI) to 5s, as shown in Supplementary Figure S1(a) and (b). 55 subjects (age = 73±6.9 years, 15 males) were enrolled and scanned on a 3T Siemens Prisma scanner (Siemens Healthcare, Erlangen, Germany) with a 32-channel head coil. All subjects provided written informed consent according to a protocol approved by the Institutional Review Board.

The conditional LDM and training were implemented with Python and Pytorch. The encoder-decoder pair was adopted from the pretrained model without finetuning. The diffusion model was trained with the data described in Supplementary Table S1. 39, 6, 10 of the whole 55 datasets were randomly selected to be the training, validation and test data, respectively. Training was performed on a Lambda cluster with NVIDIA 3090 GPU. Adam optimizer was used with initial learning rate of 2e-6 and trained for 1000 epochs. Loss function of training the denoising network were the same as described in[22],

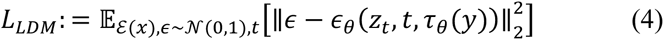

where *∈* is the noise added into the latent variable *z_t_* in diffusion step *t*.

### Model evaluation

Given the statistical properties of generative models, the generated images can vary for each sample, which enables the uncertainty measurement of the generation. We optimized the number of averaged samples by first generating 100 samples for each subject and averaging different number of samples, and then comparing the averaged image to the ground truth image to determine the best number of samples to achieve good performance. The standard deviation map of the samples was generated as a by-product to evaluate the spatial distribution of the uncertainty. To quantitatively compare the performance, we calculated the similarity (NMSE, PSNR and SSIM) between the generated M0 and the reference. To evaluate the accuracy of CBF quantification using M0 images generated from the LDM model, we calculated the same similarity metrics on CBF maps, as well as the bias in CBF values averaged in the whole brain, GM and WM regions of interest (ROI). The ROIs were segmented with SPM12 (fil.ion.ucl.ac.uk/spm/software/spm12/). To further validate whether our method complies with the physics model, we fitted the T1 maps from the generated M0 images and the control images which were compared with those fitted with acquired M0 and control images.

### Application of LDM to ADNI dataset

The conditional LDM trained on the in-house dataset was applied to the ADNI-3 dataset without finetuning. For ADNI-3 Siemens dataset, 10 pairs of control and label images were acquired for each subject. All control images were averaged and used as the condition in the LDM to generate M0 images. As the intensity of the MRI images may vary across different sites, the data were normalized before they were input to the network[33] using a histogram matching method. Specifically, the histogram of the training data was averaged and compared to the averaged values for the GM and WM regions to determine the percentile on the histogram to be used as the landmark for normalization. The average of these two landmark values were then used to normalize all ADNI data from different sites. The process of normalization can be described by the following equation:

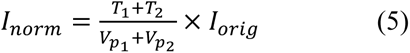

where *T*_1_ and *T*_2_ are learned landmark values for GM and WM, and 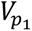 and 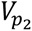 are the values of percentiles *p*_1_ and *p*_2_ on the histogram of the image to be normalized. This scale factor was calculated and saved for each individual data. After the generation of the M0 image, this scale factor was used to rescale the generated M0 back to the original scale.

For each subject in the ADNI-3 dataset, a T1-weighted structural MRI was acquired along with ASL with a resolution of 1×1×1mm^3^. In order to perform regional analysis, the generated M0 images and perfusion images were first coregistered to the T1 image and then normalized to the Montreal Neurological Institute (MNI) template space using SPM12. CBF maps were calculated and averaged for each diagnostic group, including cognitive normal (CN), mild cognitive impairment (MCI), AD and subjective memory complaints (SMC). Automated anatomical labelling (AAL) template was used to get regional CBF values within each brain regions.

### Regional analyses

Statistical analyses were performed to compare regional differences among the 4 groups, as well as the difference between GE data and generated Siemens data. A generalized linear model was used to include an interaction term between diagnostic groups and scanner types. The interaction term represented the difference between scanners of the difference among the 4 groups. A non-statistically significant interaction test indicated no statistically significant differences in CBF variation across NC, SMC, MCI and AD groups between generated (Siemens) and acquired (GE) data. Since the data did not follow the normal distribution, we conducted Wilcoxon ranking score transformations, thus all statistical tests conducted by comparing the ranking score (median) instead of mean. Model integrity was inspected by residual plots. For exploratory trajectory slope comparison, we used a mixed effect model with autoregressive covariance structure. The mixed model included all two-way interactions and a three-way interaction between diagnostic groups, scanner types and age. Contrast tests were used to estimate the slope difference between scanner types at different diagnostic groups. Since the mixed effect model is used to test on the comparison for the random slopes, the normality violation is more tolerable[34]. In this study, we did not adjust the α level for multiple comparison. As we hypothesized a similarity between generated and acquired images, a statistically non-significance would be the “positive” finding. Once α level been penalized, it will inflate the chance of claiming no-difference (the positive finding). SAS 9.4 was used for statistical analyses.

### Machine Learning analysis for binary classification

Three machine learning (ML) algorithms were used to build classifiers: Random Forest (RF), Real AdaBoost[25], and Elastic Net. RF and AdaBoost are considered as non-parametric approaches while Elastic Net is considered as parametric approach in case of strong linear predictors. The hyperparameter setting for RF was based on grid search using an interval of (5, 10, 25, 50, 100) number of variables to enter and 200 to 1000 trees by an interval of 100. Other hyperparameters were set as maximal depth of 50, leaf size of 5. The optimized hyperparameter was selected by minimizing the out of bag misclassification rate. For AdaBoost, since it is more efficient, only 25 trees were built with a depth of 3 as recommended by [35]. For Elastic Net, the hyperparameter tuning was conducted using L2 value between 0 to 1 via a grid search to minimize predicted residual sum of squares (CVPRESS). For RF and AdaBoost, Gini impurity index was used as the loss function. Loh method[36] with intensive Chi-square computing was used as for variable selection. Cross validation predicted residual sum of squares (CVPRESS) was used for Elastic Net to select candidate predictor as well as the final model. For imbalanced outcome, prior correction as described by[37] was used. For all 3 classifiers, 10-fold cross validation was used to evaluate model performance. The full dataset was equally divided into 10 folds. We re-iterated the learning process 10 times and applied the classifier to each of the testing samples. Thus, each study sample served as an independent testing case once. Receiver Operating Characteristic (ROC) curve was constructed using the predicted probability from 10 testing datasets combined and the area under the curve (AUC) with 95% confidence interval was used to assess prediction accuracy. SAS Enterprise Miner 15.1: High-Performance procedures were used for machine learning.

## CONCLUSION

In conclusion, we developed a conditional latent diffusion model for the generation of missing M0 image based on the control image in ASL acquisition, allowing CBF quantification for the ADNI-3 dataset. This methodology can be potentially applied to other large-scale and multi-site medical imaging research with missing data.

## Author Contributions

Q.S. and D.J.W. contributed to conception and design of the study. Q.S. contributed to collection of data, coding and training of the model, evaluation, data processing. N.C. contributed to additional validation data. H.K. contributed to selection of model. S.C. performed statistical analyses. J.R. conducted biological interpretations of the results. All authors contributed to the draft of the manuscript.

## Data Availability

All data produced in the present study are available upon request to the authors.

## Acknowledgements

This work was supported in part by the U.S. National Institute of Health under Grant R01-NS114382, R01-EB032169, R01-EB028297, R01-NS102220, U19-AG065169 and S10-OD032285.

## Data Availability

The analysis data is publicly available at (adni.loni.usc.edu), code for this paper will be available upon publication.

---

**Figure S1.**
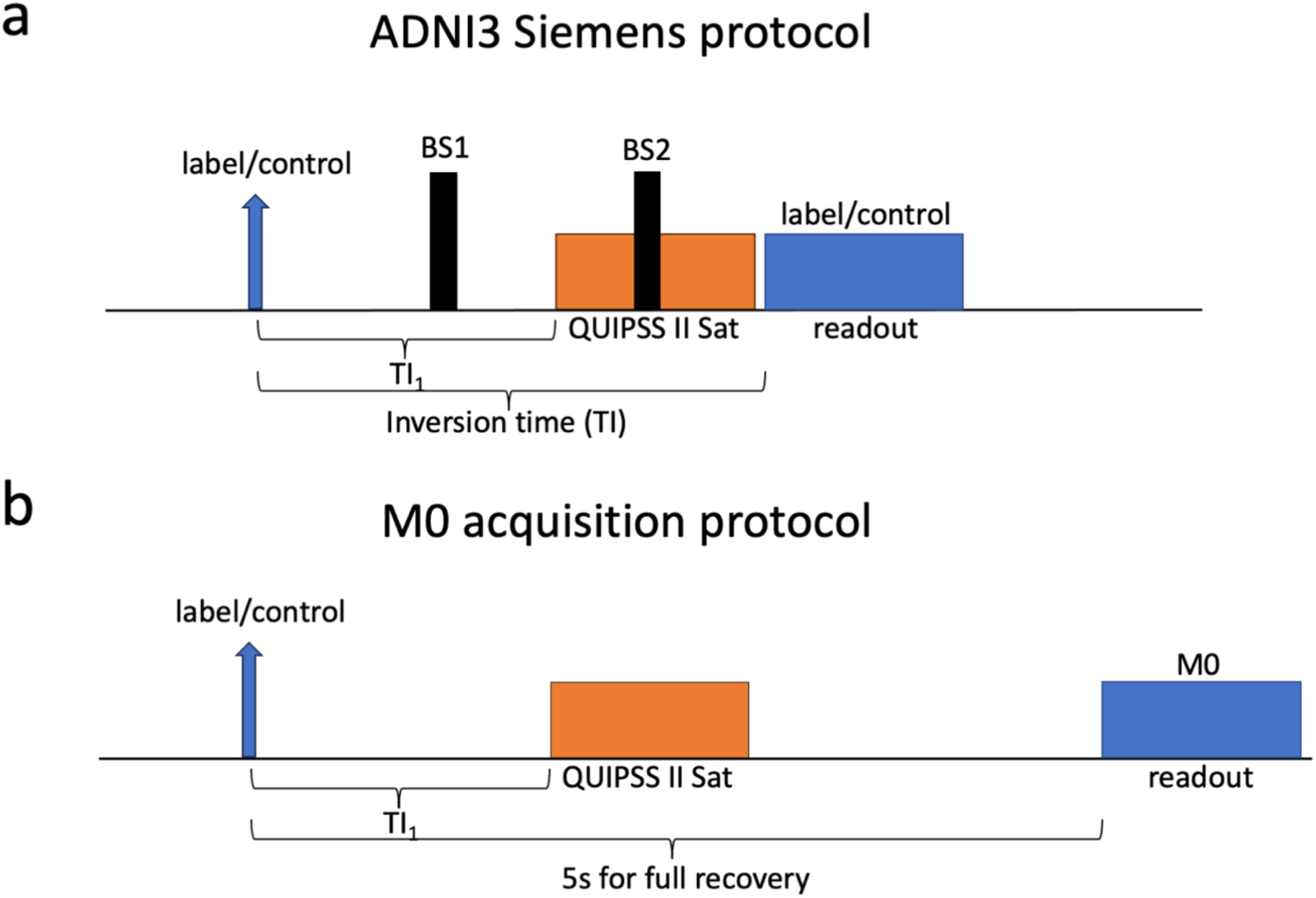
Sequence diagram of the PASL used in the ADNI protocol for Siemens scanners. a. The original product sequence to acquire control/label pairs for the PASL. b. Modified sequence to acquire M0. The background suppression pulses are disabled, and the inversion time is prolonged from 2s and 5s to enable full recovery of the longitudinal magnetization (Mz).

**Figure S2.**
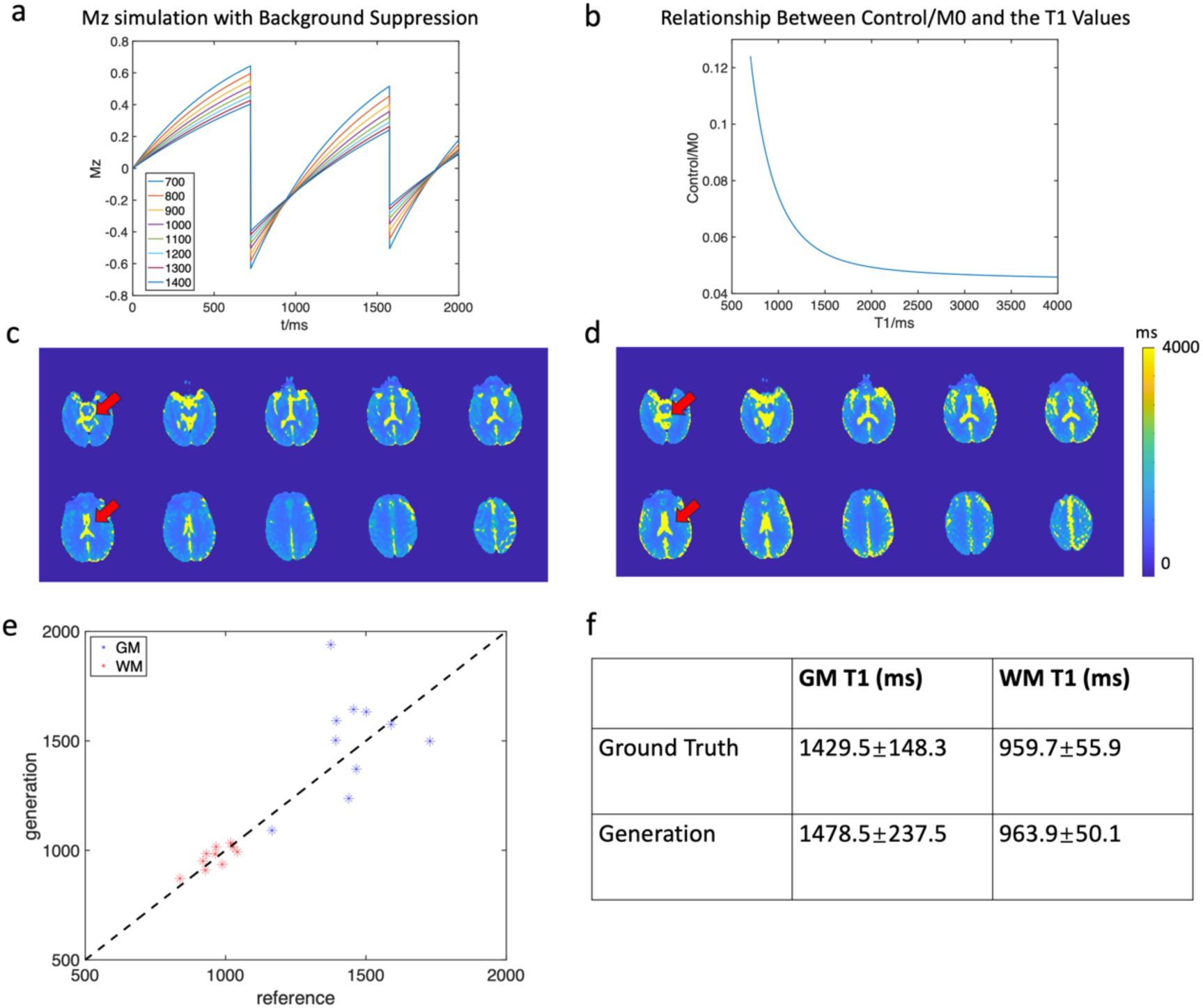
Results for T1 evaluation. a. Bloch simulation results for background suppression of various T1 values. b. Illustration of the relationship of Mz/M0 with different T1 values. c. fitted T1 map with real M0 and control images. d. fitted T1 map with generated M0 and real control images. e. scatter plot of gray matter and white matter T1 values of real and generated T1 maps. f. summary of the GM and WM values of real and generated T1 maps of all subjects.

**Figure S3.**
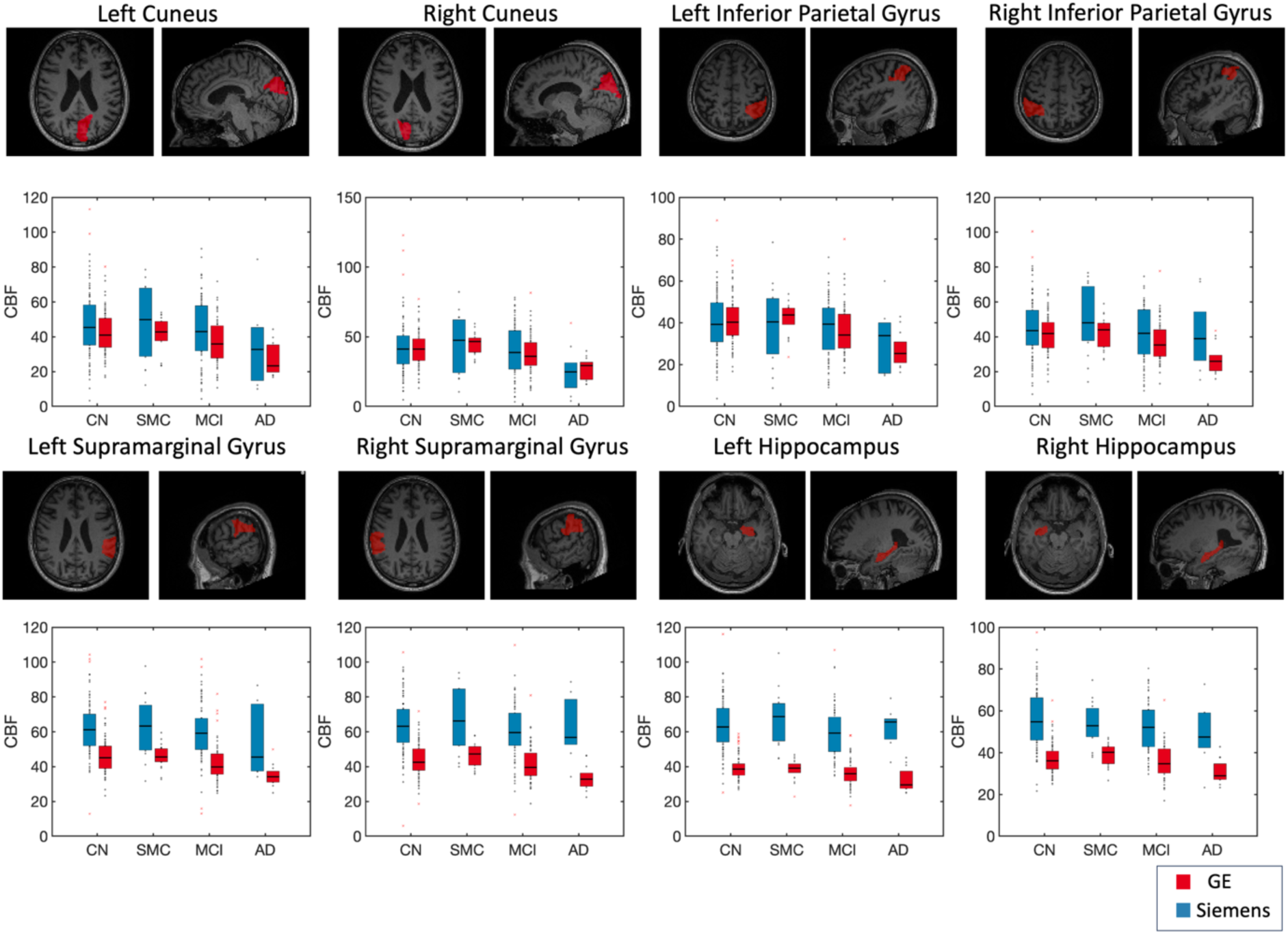
Regional analysis of more ROIs including cuneus, inferior parietal gyrus, supramarginal gyrus and hippocampus. Overall, GE and Siemens data show similar trends in different groups of subjects. Siemens data show large variance across subjects than GE data.

**Figure S4.**
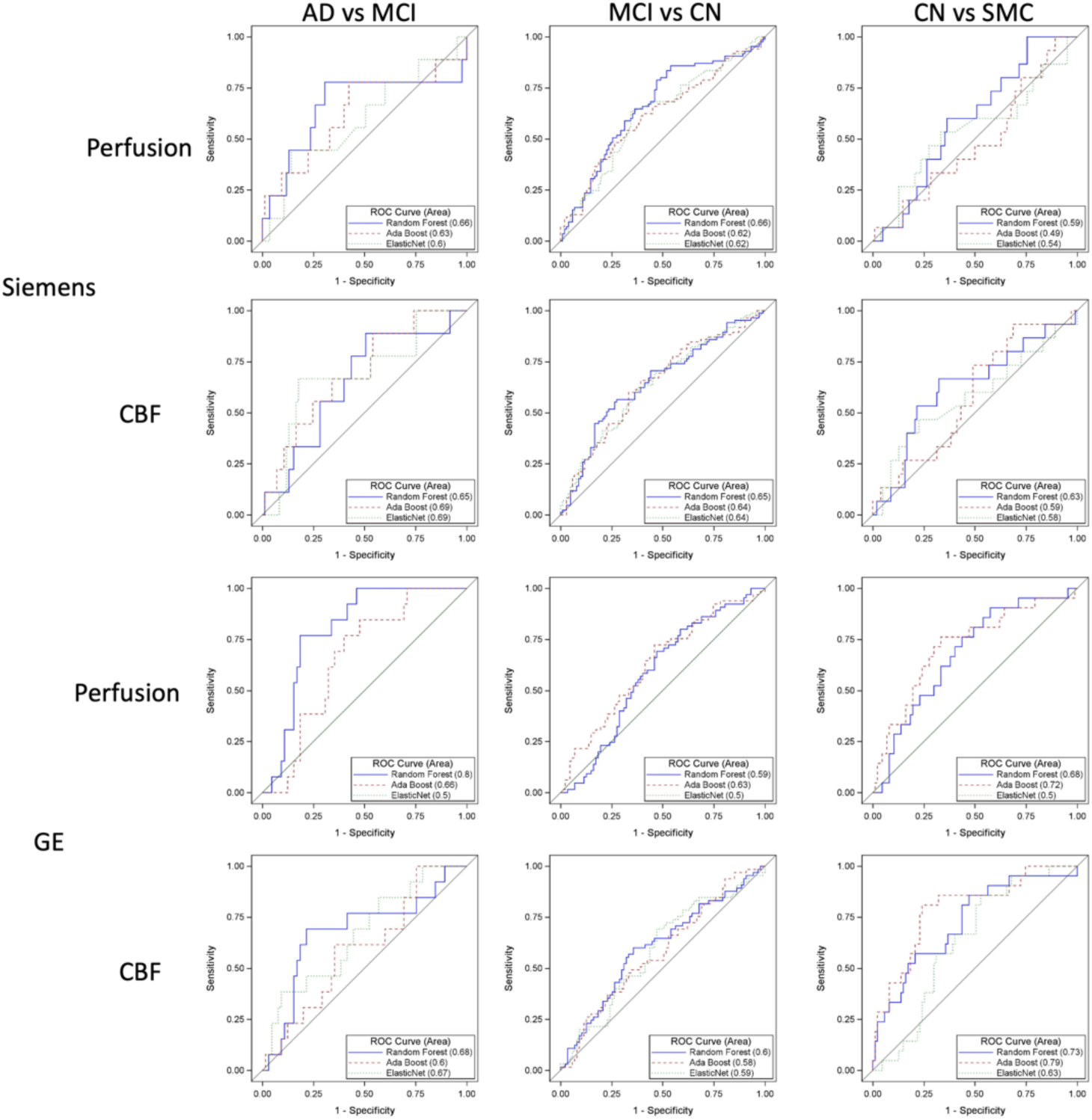
Trajectory analysis for the longitudinal data. The trend of regional CBF with age is showed in four groups of subjects. In most of the regions and groups, the trend is similar between GE and Siemens data.

**Figure S5.**
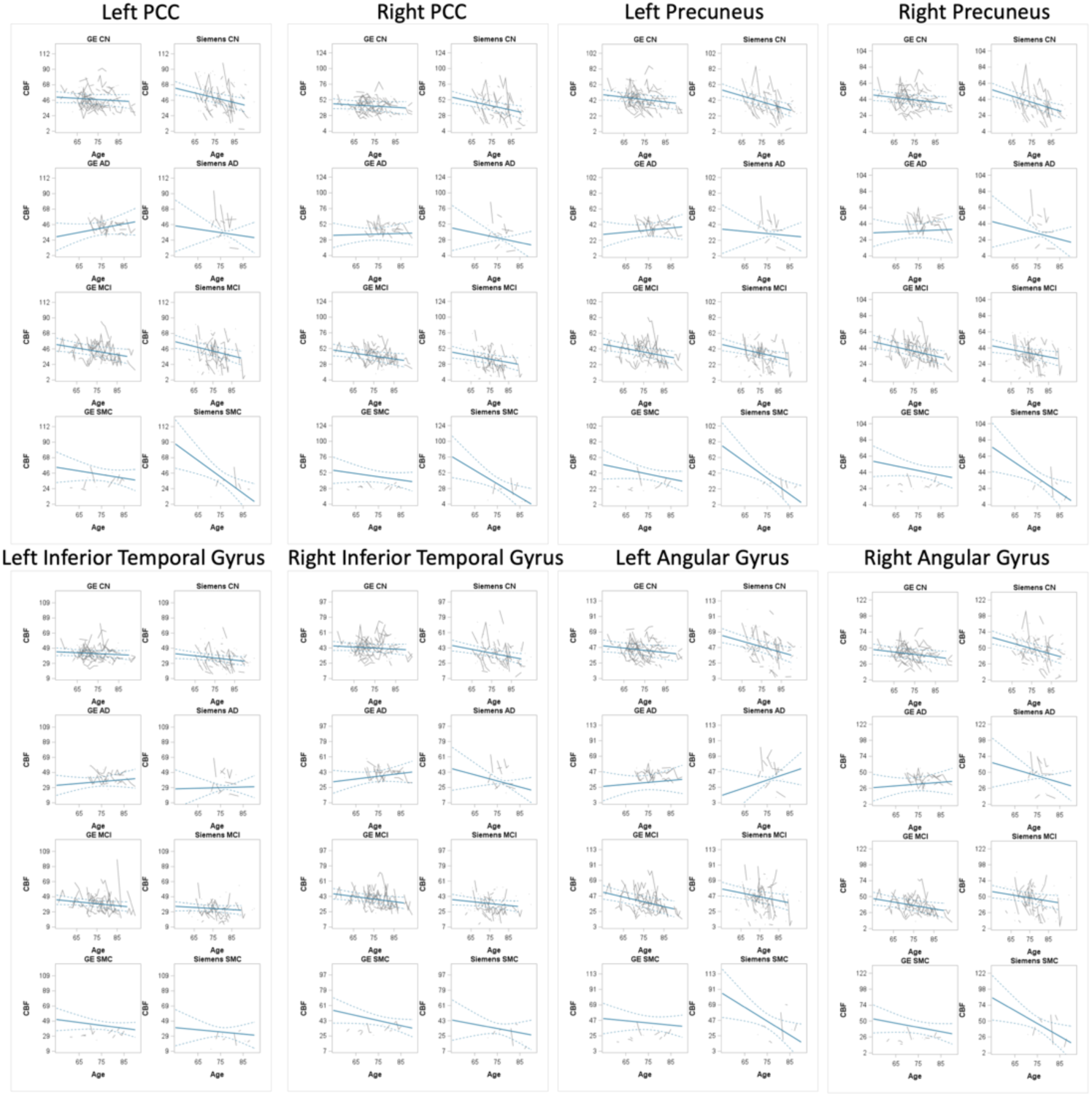
Performance of machine learning classification between AD and MCI, MCI and CN, CN and SMC. Overall, the performance is lower compared to the classification between AD and CN. Performance using CBF as features are better than the performance using perfusion as features.

## Notes

### Competing Interest Statement

The authors have declared no competing interest.

### Author Declarations

The IRB of University of Southern California gave ethical approval for this work.

