## Supplementary Table S1 for "Diffusion model enables quantitative CBF analysis of Alzheimer’s Disease"

Table S1 Demographic information for in-house dataset with paired data of ASL

| Vendor | Siemens |
| --- | --- |
| N subjects | 55 |
| Age (years) | 73±7 |
| Gender | 40 females |
| Imaging protocol | PASL |

Table S2 Demographic information for subjects (Baseline)

| Vendor | GE | Siemens |
| --- | --- | --- |
| N subjects | 186 | 211 |
| Age (years) | 73.6±7.3 | 76.2±7.1 |
| Gender | 91 females | 111 females |
| Group | CN=87; SMC=21;  MCI=65; AD=13. | CN=102; SMC=15;  MCI=85; AD=9. |
| Imaging protocol | pCASL | PASL |

Table S3 Interaction between diagnosis group and scanner types(included in the excel file)

Table S4 Results of machine learning classification with either CBF or perfusion as features. Machine learning methods include Ada Boost, Elastic Net and Random Forest.

| Machine Learning Method | Comparative groups | Vendor | AUC from CBF | AUC from Perfusion |
| --- | --- | --- | --- | --- |
| Ada Boost | MCI_CN | Siemens | 0.64 95% CI: (0.56, 0.72) | 0.62 95% CI: (0.53, 0.7) |
| Elastic Net | MCI_CN | Siemens | 0.64 95% CI: (0.56, 0.72) | 0.62 95% CI: (0.54, 0.7) |
| Random Forest | MCI_CN | Siemens | 0.65 95% CI: (0.57, 0.73) | 0.66 95% CI: (0.58, 0.74) |
| Ada Boost | SMC_CN | Siemens | 0.58 95% CI: (0.44, 0.73) | 0.48 95% CI: (0.33, 0.64) |
| Elastic Net | SMC_CN | Siemens | 0.58 95% CI: (0.4, 0.76) | 0.54 95% CI: (0.36, 0.72) |
| Random Forest | SMC_CN | Siemens | 0.63 95% CI: (0.46, 0.79) | 0.59 95% CI: (0.46, 0.73) |
| Ada Boost | AD_CN | Siemens | **0.75 95% CI: (0.62, 0.89)** | 0.67 95% CI: (0.45, 0.9) |
| Elastic Net | AD_CN | Siemens | 0.71 95% CI: (0.55, 0.87) | 0.61 95% CI: (0.39, 0.83) |
| Random Forest | AD_CN | Siemens | 0.68 95% CI: (0.5, 0.85) | 0.68 95% CI: (0.46, 0.89) |
| Ada Boost | AD_MCI | Siemens | 0.69 95% CI: (0.52, 0.87) | 0.63 95% CI: (0.39, 0.87) |
| Elastic Net | AD_MCI | Siemens | 0.69 95% CI: (0.49, 0.88) | 0.6 95% CI: (0.38, 0.82) |
| Random Forest | AD_MCI | Siemens | 0.65 95% CI: (0.47, 0.84) | 0.66 95% CI: (0.41, 0.91) |
| Ada Boost | MCI_CN | GE | 0.58 95% CI: (0.49, 0.67) | 0.63 95% CI: (0.54, 0.72) |
| Elastic Net | MCI_CN | GE | 0.59 95% CI: (0.5, 0.68) | 0.5 95% CI: (0.5, 0.5) |
| Random Forest | MCI_CN | GE | 0.6 95% CI: (0.51, 0.69) | 0.59 95% CI: (0.5, 0.68) |
| Ada Boost | SMC_CN | GE | 0.79 95% CI: (0.68, 0.9) | 0.72 95% CI: (0.59, 0.85) |
| Elastic Net | SMC_CN | GE | 0.63 95% CI: (0.52, 0.75) | 0.5 95% CI: (0.5, 0.5) |
| Random Forest | SMC_CN | GE | 0.73 95% CI: (0.6, 0.85) | 0.68 95% CI: (0.56, 0.8) |
| Ada Boost | AD_CN | GE | **0.9 95% CI: (0.83, 0.98)** | 0.81 95% CI: (0.72, 0.91) |
| Elastic Net | AD_CN | GE | 0.84 95% CI: (0.74, 0.94) | 0.5 95% CI: (0.5, 0.5) |
| Random Forest | AD_CN | GE | 0.8 95% CI: (0.64, 0.96) | 0.82 95% CI: (0.71, 0.93) |
| Ada Boost | AD_MCI | GE | 0.6 95% CI: (0.43, 0.76) | 0.66 95% CI: (0.53, 0.8) |
| Elastic Net | AD_MCI | GE | 0.67 95% CI: (0.5, 0.83) | 0.5 95% CI: (0.5, 0.5) |
| Random Forest | AD_MCI | GE | 0.68 95% CI: (0.5, 0.86) | 0.8 95% CI: (0.7, 0.9) |

Table S5 Demographic information for subjects (Longitudinal)

| Vendor | GE | Siemens |
| --- | --- | --- |
| N subjects (1 timepoint) | 62 | 115 |
| N subjects (2 timepoints) | 38 | 65 |
| N subjects (3 timepoints) | 53 | 25 |
| N subjects (4 timepoints) | 21 | 5 |
| N subjects (5 timepoints) | 7 | 1 |
| N subjects (6 timepoints) | 5 | 0 |
| Total | 186 | 211 |

Table S6 Slope difference comparison between GE and Siemens in longitudinal dataset. (included in the excel file)
